# Evaluating Progress in Automatic Chest X-Ray Radiology Report Generation

**DOI:** 10.1101/2022.08.30.22279318

**Authors:** Feiyang Yu, Mark Endo, Rayan Krishnan, Ian Pan, Andy Tsai, Eduardo Pontes Reis, Eduardo Kaiser Ururahy Nunes Fonseca, Henrique Min Ho Lee, Zahra Shakeri Hossein Abad, Andrew Y. Ng, Curtis P. Langlotz, Vasantha Kumar Venugopal, Pranav Rajpurkar

**Author notes:** Corresponding author: Pranav Rajpurkar, PhD. These authors contributed equally: Feiyang Yu, Mark Endo, Rayan Krishnan.

## Abstract

The application of AI to medical image interpretation tasks has largely been limited to the identification of a handful of individual pathologies. In contrast, the generation of complete narrative radiology reports more closely matches how radiologists communicate diagnostic information in clinical workflows. Recent progress in artificial intelligence (AI) on vision-language tasks has enabled the possibility of generating high-quality radiology reports from medical images. Automated metrics to evaluate the quality of generated reports attempt to capture overlap in the language or clinical entities between a machine-generated report and a radiologist-generated report. In this study, we quantitatively examine the correlation between automated metrics and the scoring of reports by radiologists. We analyze failure modes of the metrics, namely the types of information the metrics do not capture, to understand when to choose particular metrics and how to interpret metric scores. We propose a composite metric, called RadCliQ, that we find is able to rank the quality of reports similarly to radiologists and better than existing metrics. Lastly, we measure the performance of state-of-the-art report generation approaches using the investigated metrics. We expect that our work can guide both the evaluation and the development of report generation systems that can generate reports from medical images approaching the level of radiologists.

## Introduction

Artificial Intelligence (AI) has been making great strides in tasks that require expert knowledge, such as playing Go^1–4^, writing code^5,6^, and driving vehicles^7,8^. In the medical domain, AI has reached similar exciting milestones^9^, including the effective prediction of 3D protein structures^10,11^. Enabled by the rapidly evolving imaging and computer vision technologies, AI also has made formidable progress on image interpretation tasks, including chest X-ray interpretation. However, the application of AI to image interpretation tasks has often been limited to the identification of a handful of individual pathologies^12–14^, representing an over-simplification of the image interpretation task. In contrast, the generation of complete narrative radiology reports^15–20^ moves past that simplification and matches up to how radiologists communicate diagnostic information: the narrative report allows for highly diverse and nuanced findings, including association of findings with anatomic location, and expressions of uncertainty. Although the generation of radiology reports in their full complexity would signify a tremendous achievement for AI, the task remains far from solved. Our work aims to tackle one of the most important bottlenecks for progress: *the limited ability to meaningfully measure progress on the report generation task*.

Automatically measuring the quality of generated radiology reports is challenging. Most prior works have relied on a set of automated metrics inspired by similar setups in natural language generation, where radiology report text is treated as generic text^21^. However, unlike generic text, radiology reports involve complex, domain-specific knowledge and critically depend on factual correctness. Even metrics that were designed to evaluate the correctness of radiology information by capturing domain-specific concepts do not align with radiologists^22^. Therefore, improvement on existing metrics may not produce clinically meaningful progress or indicate the direction for further progress. This fundamental bottleneck hinders understanding of the quality of report generation methods thereby impeding work toward improvement of existing methods. We seek to remove this bottleneck by developing meaningful measures of progress in radiology report generation. The answer to this question is imperative to understanding which metrics can guide us towards generating reports that are clinically indistinguishable from those generated by radiologists.

In this study, we quantitatively examine the correlation between automated metrics and the scoring of reports by radiologists. We propose a new automatic metric which computes the overlap in clinical entities and relations between a machine-generated report and a radiologist-generated report, called RadGraph^23^ F1. We develop a methodology to predict a radiologist-determined error score from a combination of automated metrics, called RadCliQ. We analyze failure modes of the metrics, namely the types of information the metrics do not capture, to understand when to choose particular metrics and how to interpret metric scores. Lastly, we measure the performance of state-of-the-art report generation models using the investigated metrics. The result is a quantitative understanding of radiology report generation metrics and clear guidance for metric selection to guide future research on automated chest X-ray interpretation. This work is also broadly applicable to medical imaging interpretation and narrative report generation in other domains.

## Results

### Quantitative investigation of alignment between automated metrics and radiologists

We study whether there is high alignment between automated metric and radiologist scores assigned to radiology reports. Given a test report from the MIMIC-CXR^24–26^ test set, we select a series of candidate reports from the MIMIC-CXR train set that score highly according to various metrics. We choose this set of reasonably accurate reports so we can study their quality with more precision. Next, we have radiologists score how well the candidates match the test report. We can then analyze the alignment between radiologist and metric scores and determine how correlated different metrics are with radiologists. We select a candidate report by finding the test report’s *metric-oracle*: the highest-scoring report from the MIMIC-CXR training set with respect to a particular metric. Since the metric-oracle reports are the best possible retrievals according to a metric, they represent the theoretical best performance achievable by methods that retrieve reports from the training corpus to describe input X-ray images. Although the metric-oracle approach is not a viable clinical method for reporting, it is useful as part of a framework to study report metrics.

### Metric-oracle reports

We constructed metric-oracle reports for four metrics. These include BLEU^27^, BERTScore^28^, CheXbert vector similarity (s_emb)^13^ and a novel metric RadGraph^23^ F1. BLUE and BERTScore are general natural language metrics for measuring the similarity between machine-generated and human-generated texts. BLEU computes n-gram overlap and is representative for the family of text overlap based natural language generation metrics such as CIDEr^29^, METEOR^30^ and ROUGE^31^. BERTScore has been proposed for capturing contextual similarity beyond exact textual matches. CheXbert vector similarity and RadGraph F1 are metrics designed to measure the correctness of clinical information. CheXbert vector similarity computes the cosine similarity between the indicator vectors of 14 pathologies that the CheXbert automatic labeler extracts from machine-generated and human-generated radiology reports. It is designed to evaluate radiology specific information but its evaluation is limited to 14 pathologies. To address this limitation, we propose the use of the knowledge graph of the report to represent arbitrarily diverse radiology specific information. We design a novel metric, RadGraph F1, that computes the overlap in clinical entities and relations that RadGraph extracts from machine- and human-generated reports. The four metrics are detailed in the Methods section.

For every test report, we generated the matching metric-oracle report by selecting the highest scoring report, according to each of the four investigated metrics, from the training set. We specifically used the impression section of the report. As an example of our setup, for the test report of “*No acute cardiopulmonary process. Bilateral low lung volumes with crowding of bronchovascular markings and bibasilar atelectasis*,” the metric-oracle retrieved with respect to BERTScore was: “*No acute cardiopulmonary process. Low lung volumes and bibasilar atelectasis*,” while the metric-oracle retrieved with respect to RadGraph F1 was: “*No acute cardiopulmonary process. Bilateral low lung volumes*,” as shown in Fig. 2(a).

**Fig. 1:**
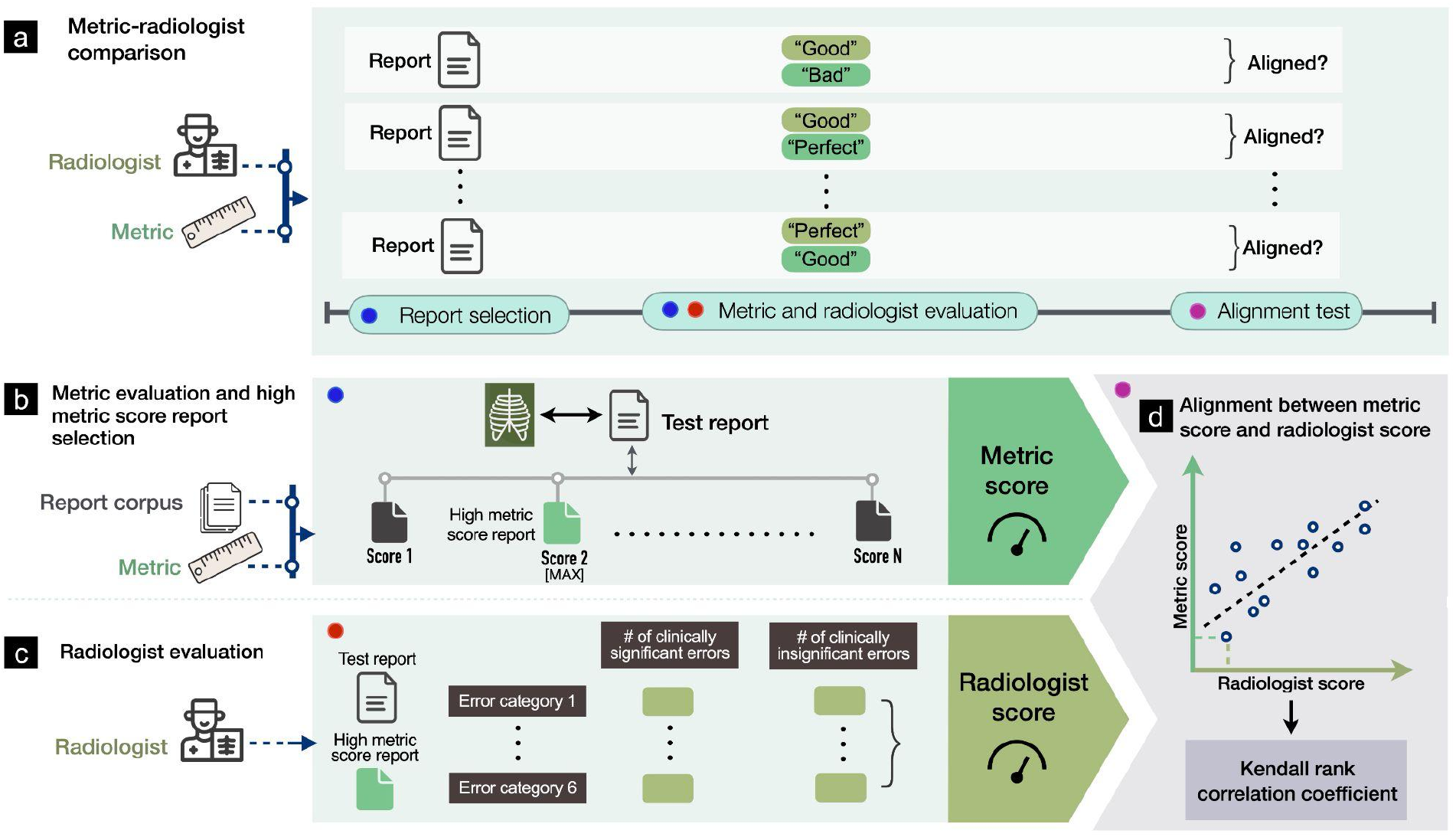
**a**, Experimental design for selecting radiology reports and comparing metrics and radiologists in evaluating reports. **b**, Given a test report, selecting the report with the highest metric score from the training report corpus with respect to the test report and a particular metric. **c**, Conducting radiologist evaluation on the high metric score report relative to the test report, where radiologists identify the number of clinically significant and insignificant errors in the high metric score report across six error categories. **d**, Determining the alignment between metric scores and radiologist scores assigned to the same reports using the Kendall rank correlation coefficient.

**Fig. 2:**
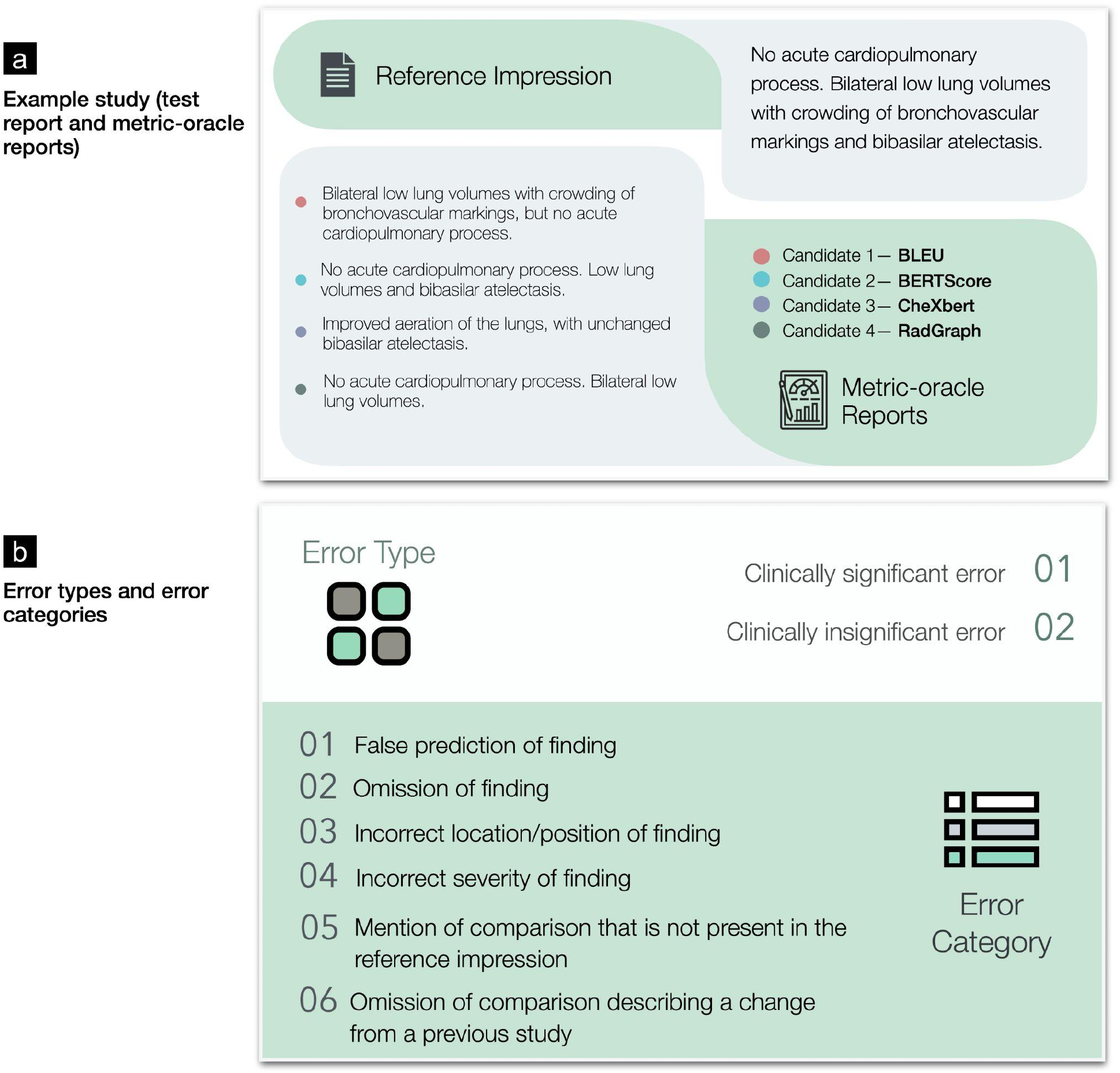
**a**, Example study of a test report and four metric-oracle reports corresponding to BLEU, BERTScore, CheXbert vector similarity and RadGraph F1 that radiologists evaluate to identify errors. **b**, Two error types and six error categories that radiologists identify for each pair of test report and metric-oracle report.

### Radiologist evaluation study design

In our experimental study design, six board certified radiologists scored the number of errors that various metric-oracle reports make compared to the test report. Radiologists categorized errors as significant or insignificant. Radiologists subtyped every error into the following six categories: 1) false prediction of finding (i.e., false positive) 2) omission of finding (i.e., false negative) 3) incorrect location/position of finding 4) incorrect severity of finding 5) mention of comparison that is not present in the reference impression, and 6) omission of comparison describing a change from a previous study. We sampled 50 studies randomly from the MIMIC-CXR test set. The ordering of metrics that the metric-oracle reports correspond to was shuffled for every study. The error types and error categories are summarized in Fig. 2(b). The instructions and interface presented to radiologists can be seen in Supplementary Fig. 1.

### Alignment between automated metrics and radiologists

We first quantify metric-radiologist alignment using the Kendall rank correlation coefficient (tau-b) between metric scores and number of radiologist-reported errors in the reports. We determine the metric-radiologist alignment from metric-oracle generations from 50 chosen studies on both a total error and significant error level. We find that BERTScore and RadGraph F1 are the metrics with the two highest alignments with radiologists. Specifically, BERTScore has a tau value of 0.500 [95% CI 0.497 0.503] for total number of errors and 0.496 [95% CI 0.493 0.498] for significant errors. RadGraph has a tau value of 0.463 [95% CI 0.460 0.465] for total number of errors and 0.459 [95% CI 0.456 0.461] for significant errors. We find that BLEU is the third best metric under this evaluation with a 0.459 [95% CI 0.456 0.462] tau value for total number of errors and 0.445 [95% CI 0.442 0.448] for significant errors. Lastly, CheXbert vector similarity has the worst alignment with a tau value of 0.457 [95% CI 0.454 0.459] for total errors and 0.418 [95% CI 0.416 0.421] for significant errors. From these results, we see that BERTScore, RadGraph, and BLEU are the metrics with closest alignment to radiologists. CheXbert has alignment with radiologists but is less concordant than the previously mentioned metrics. The metric-radiologist alignment graphs are shown in Fig. 3.

**Fig. 3:**
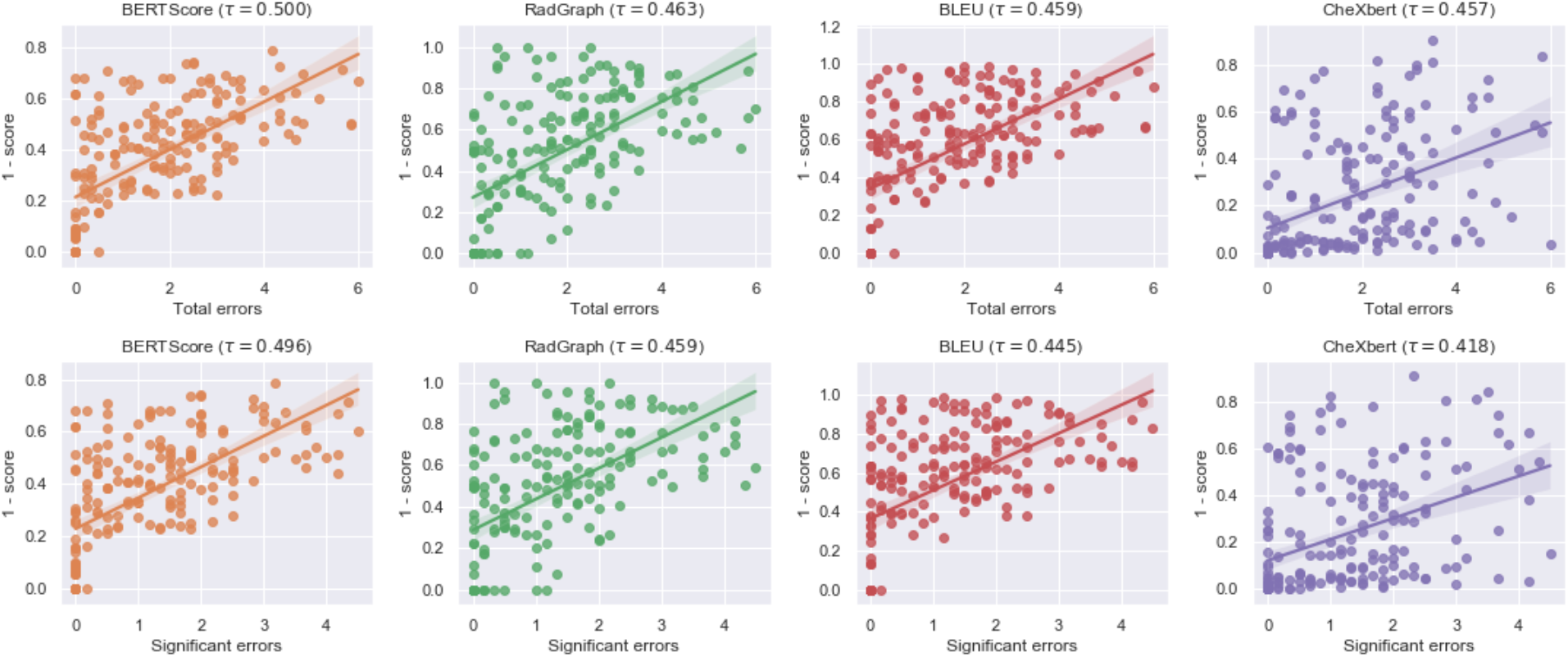
Scatter plots and correlations between metric scores and radiologist scores of four metric-oracle generations from 50 studies, where radiologist scores are represented by the total number of errors (top row) and number of clinically significant errors (bottom row) identified by the radiologists.

### Failure modes of metrics

In addition to evaluating the clinical relevance of metrics in terms of the total number of clinically significant and insignificant errors, we also examine the particular error categories of metric-oracles to develop a granular understanding of the failure modes of different metrics, as shown in Fig. 4. We use the following six error categories as described earlier:

**Fig. 4:**
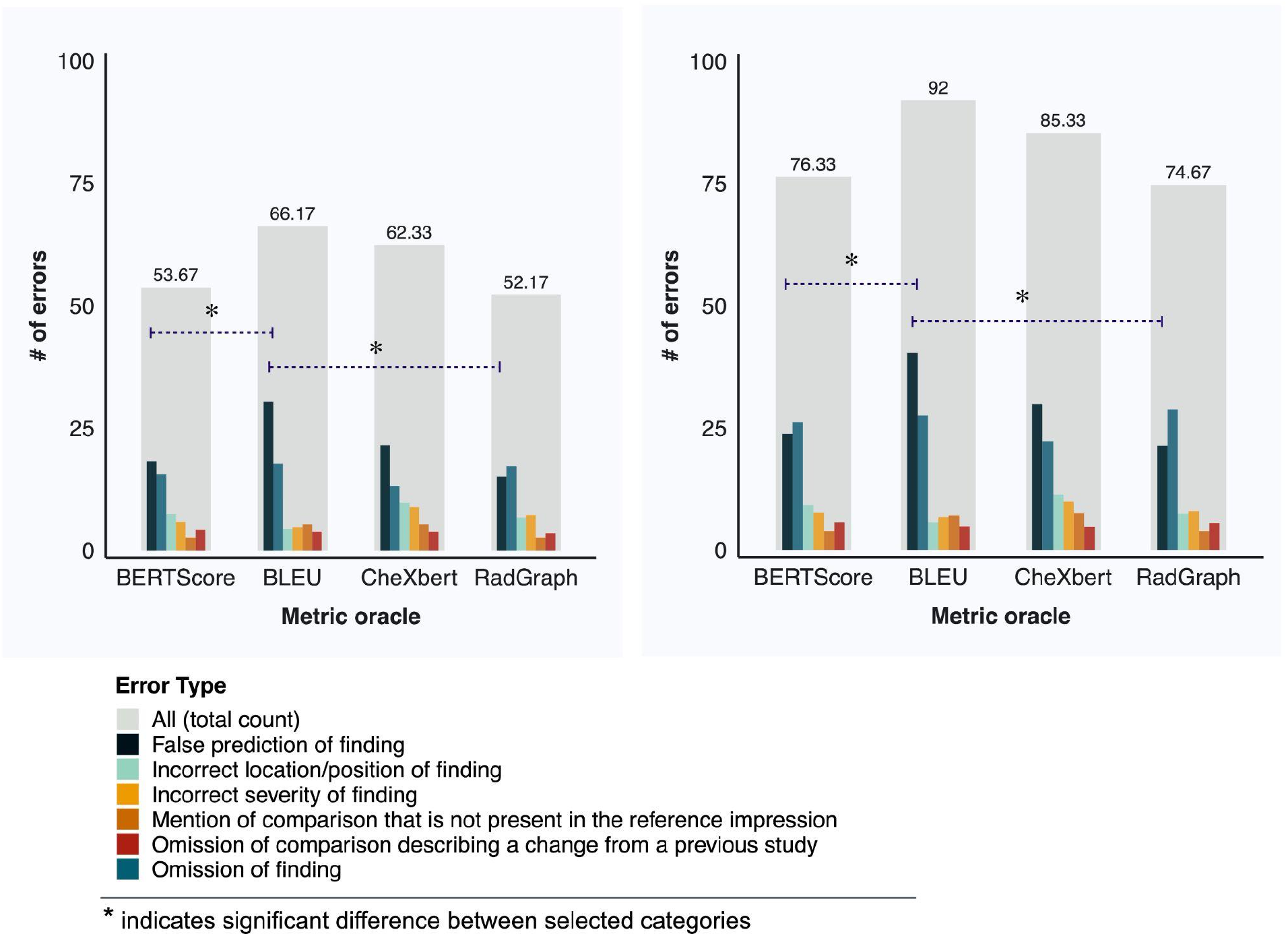
Distribution of errors across six error categories for metric-oracle reports corresponding to BERTScore, BLEU, CheXbert vector similarity and RadGraph F1, in terms of the number of clinically significant errors (left) and the total number of errors (right).

1. False prediction of finding
2. Omission of finding
3. Incorrect location/position of finding
4. Incorrect severity of finding
5. Mention of comparison that is not present in the reference impression
6. Omission of comparison describing a change from a previous study

and analyze the total number of errors and the number of clinically significant errors within each error category.

BLEU exhibits a prominent failure mode in identifying false predictions of finding in reports. Metric-oracle reports with respect to BLEU produce more false predictions of finding than BERTScore and RadGraph in terms of both the total number of errors (0.807 average number of errors per report versus 0.477 and 0.427 for BERTScore and RadGraph) and the number of clinically significant errors (0.607 average number of errors per report versus 0.363 and 0.300 for BERTScore and RadGraph). BLEU exhibits a less prominent failure mode in identifying incorrect locations/positions of finding compared with CheXbert vector similarity. Metric-oracle reports with respect to BLEU have fewer incorrect locations/positions of finding than CheXbert in terms of both the total number of errors (0.113 average number of errors per report versus 0.227 for CheXbert) and the number of clinically significant errors (0.087 average number of errors per report versus 0.193 for CheXbert). These differences are statistically significant after accounting for multiple-hypothesis testing. Metric-oracle reports of the four metrics exhibit similar behavior in the other error categories, as the differences in number of errors are not statistically significant. The raw error counts and the statistics testing results for two-sample t tests and the Benjamini-Hochberg Procedure for accounting for multiple-hypothesis testing are shown in Supplementary Table 2 and Supplementary Table 3.

### Measuring progress of prior methods in report generation

Using the four individual metrics, we evaluated the following state-of-the-art radiology report generation methods: M^2^ Trans^15^, R2Gen^16^, CXR-RePaiR^17^, WCL^18^, and CvT2DistilGPT2^19^. As a baseline, we also implemented a random radiology report generation model, which retrieves a random report from the training set for each test report. We measured the performances of metric-oracle selection models and prior models relative to the baseline in terms of percentage change in metric scores, as shown in Fig. 5(a)-(d). The performances of prior methods with respect to all metrics are statistically significantly lower than those of metric-oracle methods. With respect to BLEU, the metric-oracle selection model achieves an average score of 0.566 [95% CI 0.566 0.566], while the best prior model, M^2^ Trans, achieves 0.087 [95% CI 0.087 0.087]. With respect to RadGraph F1, the metric-oracle selection model achieves an average score of 0.677 [95% CI 0.677 0.678], while the best prior model, M^2^ Trans, achieves 0.110 [95% CI 0.110 0.111]. We also note that BLEU and RadGraph correctly rank all prior models above the random retrieval baseline, while BERTScore and CheXbert vector similarity do not. This suggests that BLEU and RadGraph work sensibly with generations of real-world report generation models that only have access to input X-ray images, while the other two metrics do not.

**Fig. 5:**
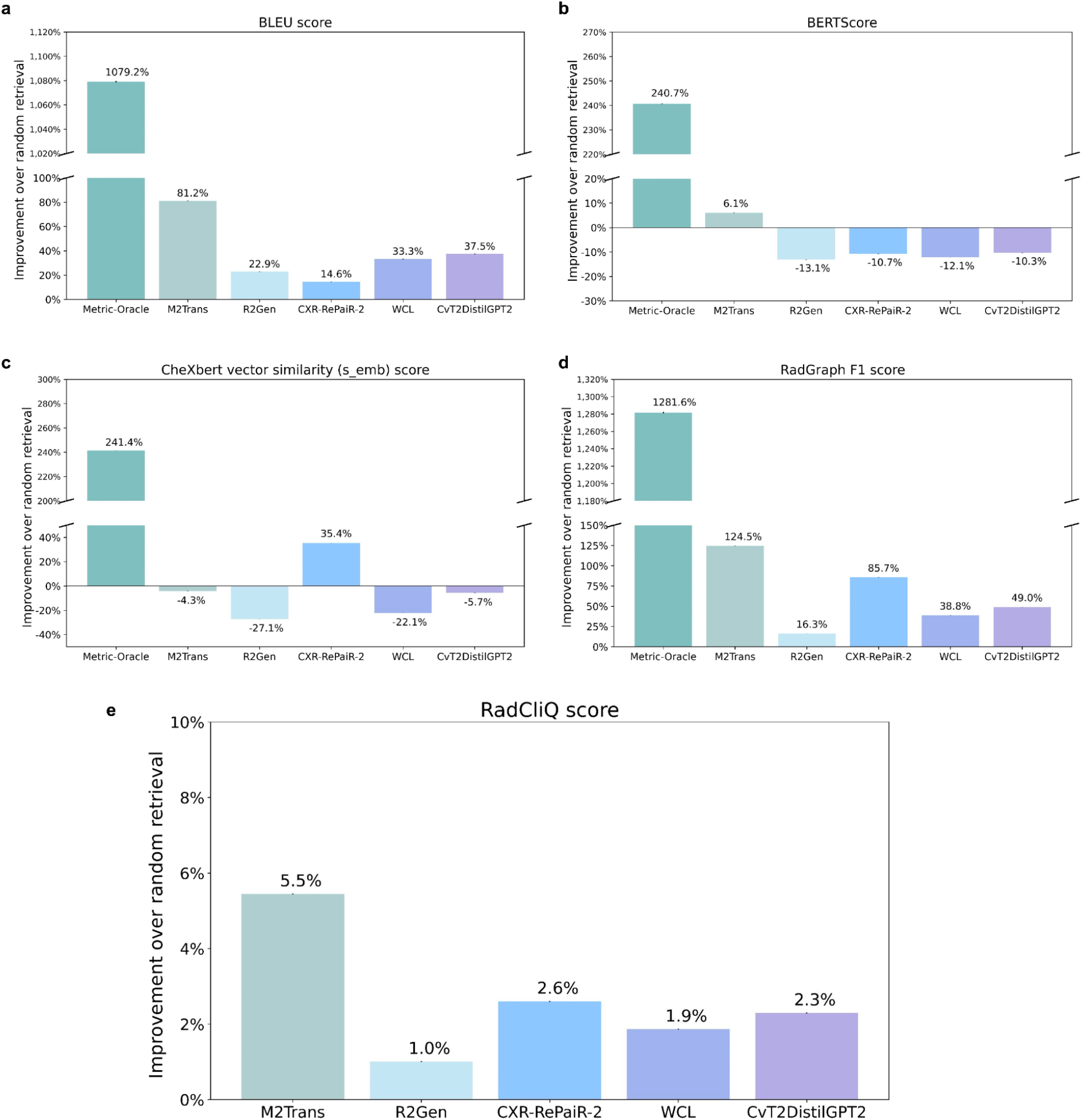
**a-d**, The percentage change in metric scores of the corresponding metric-oracle model and state-of-the-art report generation models relative to the baseline random model, for BLEU, BERTScore, CheXbert vector similarity and RadGraph F1. A higher value indicates better performance as evaluated by the metric. The raw results can be found in Supplementary Table 4. **e**, The percentage decrease in the predicted total number of errors relative to the baseline random model according to the composite metric RadCliQ. A higher value indicates better performance as evaluated by the metric. The raw results can be found in Supplementary Table 5.

### Composite metric “RadCliQ” (Radiology Report Clinical Quality)

To improve upon individual metrics, we propose a novel composite metric *RadCliQ* (Radiology Report Clinical Quality). Given that BLEU and RadGraph F1 correctly assigned higher scores to generations of prior state-of-the-art models than randomly retrieved reports, we combined BLEU and RadGraph F1 to build a composite metric. We trained a linear regression model to predict the total number of errors that radiologists would assign to a report. The model input consisted of the two metric scores computed for each report, with the scores corresponding to each metric independently normalized to have a mean of 0 and standard deviation of 1. Prediction of the trained model therefore combines evaluations of BLEU and RadGraph F1. The model was trained on the same set of 200 metric-oracle reports which were evaluated by radiologists, containing 50 metric-oracle reports corresponding to each of the four investigated metrics. The regression produced an R^2^ correlation coefficient of 0.423. The coefficients were -0.559 for BLEU and -0.526 for RadGraph F1. The intercept value for the regression model was 1.642, indicating the number of errors for a report with an average score across metrics. For each individual metric, a higher metric score indicates better report generation. Since the linear regression model was trained to predict the total number of errors in a report, the two metrics both had a negative correlation with the predicted number of errors.

As an additional statistical test, the composite metric RadCliQ had a Kendall-tau b correlation coefficient of 0.522 [95% CI 0.520 0.525] (p-value < 0.01) for the total number of errors. This indicates a statistically significant correlation between the predicted number of errors and the true number of errors in the generated reports, suggesting that the composite metric aligns with radiologists. Furthermore, RadCliQ has a stronger alignment with radiologists than any individual metric.

Using RadCliQ, we measured the performance of prior models relative to the baseline in terms of the percentage decrease in the predicted total number of errors, as shown in Fig. 5(e), where a more positive value translates to better performance. M^2^ Trans yields an 5.5% improvement over the baseline and has the best performance; CXR-RePaiR yields an 2.6% improvement and has the second best performance; CvT2DistilGPT2 yields an 2.3% improvement and has the third best performance. Here, we normalize the individual metric scores using training time statistics before computing RadCliQ.

## Discussion

The purpose of this study was to investigate how to meaningfully measure progress in radiology report generation. We studied popular existing automated metrics and also designed novel metrics, the RadGraph graph overlap metric and the composite metric RadCliQ, for report evaluation. We quantitatively determined the alignment of metrics with clinical radiologists and the reliability of metrics against specific failure modes, clarifying whether metrics meaningfully evaluate radiology reports and therefore can guide future research in report generation. We also showed that selecting the best-match report from a large corpus performs better on most metrics that the current state-of-the-art radiology report generation methods. Although the best-match method is unlikely to be clinically viable, it served as a useful tool to derive the RadCliQ composite metric developed in this study and could serve as a useful benchmark against which to evaluate report generation algorithms developed in the future.

The design of automated evaluation metrics that are aligned with manual expert evaluation has been a challenge for research in radiology report generation as well as medical report generation as a whole. Prior works have used metrics designed to improve upon n-gram matching^27–31^ or include clinical awareness^12,13,15,23,17^, such as with BLEU^27^ and CheXpert labels^12^. However, these evaluations nevertheless poorly approximate radiologists’ evaluation of reports. The expressivity of prior metrics is often restricted to a curated set of medical conditions. Therefore, the quantitative investigation of metric-radiologist alignment conducted in this study is necessary for understanding whether these metrics meaningfully evaluate reports. Prior works have investigated the alignment between metrics and human judgment^22,32^. However, to the best of our knowledge, these works pose one of two limitations for radiology report evaluation: (1) they study metric alignment with humans for general image captioning, which does not involve radiology specific terminology, a high prevalence of negation, or expert human evaluators, and (2) they do not create a leveled comparison between metrics and radiologists, where metrics and radiologists assign scores to reports in identical experimental settings, or a granular understanding of metric behavior beyond the overall metric score. Our work builds a fair comparison between general natural language and clinically aware metrics and radiologists by providing them with the same set of information that is the reports and goes beyond metric scores to examine six granular failure modes of each metric. Additionally, our work proposes a novel composite metric, RadCliQ, that aligns more strongly than any individual metric. We also show that current radiology report generation algorithms exhibit relatively low performance by all of these metrics.

To study metric-radiologist alignment, we designed *metric-oracles*: the reports selected from a large corpus with the highest metric score with respect to test reports. We had metrics and radiologists assign scores to the metric-oracles based on how well the metric-oracles match their respective test reports, and computed the alignment between metric and radiologist scores on the same reports. Pairing metric-oracles with test reports produces a narrower distribution of scores than using random reports. However, metric oracles are necessary for obtaining reliable scores from the radiologist experiment because comparisons will be sensitive to small differences in report quality. If a random report, rather than a high-scoring report, was paired with the test report, the two reports could diverge to the extent that they were difficult to compare directly. In contrast, metric oracles are comparable with test reports and therefore allow a meaningful evaluation of errors.

To generate metric-oracles, any report generation model is theoretically feasible. There are three main categories: the first generates free text based on semantics extracted from input chest X-ray images^20,33,34^; the second retrieves existing text that best matches input images from a report corpus^17,35^; and the third selects curated templates corresponding to a predefined set of abnormalities^14,36^. We chose to use retrieval-based models to generate metric-oracles because retrieval from a training report corpus produces a controlled output space, instead of an unpredictable one produced by models that generate free text. Retrieval-based models also improve upon templating-based models in terms of flexibility and generalizability because the report corpus better captures real-world occurring conditions, combinations of conditions, and uncertainty. Furthermore, retrieval-based metric-oracle models outperformed existing report generation methods by a large margin.

By investigating the different categories of errors that radiologists identified in metric-oracle reports, we also uncovered specific metric failure modes which valuably inform the choice of metrics and interpretation of metric scores for evaluating generated reports. We find that BLEU performs worse than BERTScore and RadGraph in evaluating false prediction of finding. Yet, BLEU performs better than CheXbert vector similarity in evaluating incorrect position/location of finding. Therefore, BERTScore and RadGraph, which offer the strongest radiologist-alignment, also have better overall reliability against failure modes.

Using the individual metrics and RadCliQ, we also measured the progress of prior state-of-the-art models. We identify M^2^ Trans as the best model with respect to all individual metrics and RadCliQ except CheXbert. We also find a statistically significant performance gap between prior models and metric-oracle models, which represent the theoretical performance ceiling of retrieval-based methods on MIMIC-CXR. This gap suggests that prior models in report generation still have significant room for improvement in creating high-quality reports that are useful to radiologists.

In addition, we observed that BLEU and RadGraph correctly rank prior models above random retrieval while BERTScore and CheXbert do not, even though BERTScore has the strongest alignment with radiologists in evaluating metric-oracle reports. This discrepancy may be attributed to two factors. First, there is a shift in report quality from metric-oracles to real generations, and metrics may exhibit different behaviors with reports of lower quality. Second, the real-world models generated reports based on only the X-ray images. The images may contain different semantics than that described in the corresponding test reports. For expressing the same semantics, the models also have numerous ways to formulate a report. These variations can explain BERTScore’s suboptimal performance in evaluating prior model generations. Overall, RadGraph is the best individual metric to use for its strong alignment with radiologists, reliability across failure modes and meaningful empirical performance with real generations.

This study has several important limitations. A main limitation is the inter-observer variability in radiologist evaluation. Although the evaluation scheme–the separation of clinically significant and insignificant errors, and the six error categories–was designed to be objective and consistent across radiologist evaluation, the same report often received varying scores between radiologists, a common occurrence in experiments that employ subjective ratings from clinicians. This suggests a potential limitation of the evaluation scheme used, but may also present an intrinsic problem with objective evaluation of radiology reports. Another limitation is the coverage of metrics. Although a variety of general and clinical natural language metrics are investigated, there exist other metrics in these two categories that may have different behaviors than the four investigated metrics. For instance, other text overlap based metrics are commonly used in natural language generation beyond BLEU, such as CIDEr^29^, METEOR^30^ and ROUGE^31^, which may have better or worse radiologist-alignment and reliability than BLEU in report generation.

In this study, we determined that the novel metrics RadGraph F1 and RadCliQ meaningfully measure progress in radiology report generation and hence can guide future report generation models in becoming clinically indistinguishable from radiologists. We have open-sourced the code for computing the individual metrics and RadCliQ on reports in the hope of facilitating future research in radiology report generation.

## Online Methods

### Datasets

We used the MIMIC-CXR dataset to conduct our study. The MIMIC-CXR dataset^24–26^ is a de-identified and publicly available dataset containing chest X-ray images and semi-structured radiology reports from the Beth Israel Deaconess Medical Center Emergency Department. There are 227,835 studies with 177,110 images conducted on 65,379 patients. We used the impression section of the reports. We used the recommended train/validation/test split. We pooled the train and validation splits as the training report corpus from which metric-oracles are retrieved and used the test split as the set of ground-truth reports. The training report corpus contains 185,538 studies and 371,951 images; the test set contains 2,192 studies and 5,159 images. We preprocessed the reports by filtering nan reports and extracting the impression section of reports, which contains key observations and conclusions drawn by radiologists. Throughout the study, we refer to the impression section when discussing reports.

### Advantages of metric-oracles

Using metric-oracles as the candidate reports as opposed to using other strategies such as randomly sampling reports offers two primary advantages: (1) metric-oracles are sufficiently accurate for radiologists to pinpoint specific errors and not be bogged down by candidate reports that aren’t remotely similar to the test reports; (2) metric-oracles allow us to analyze where certain metrics fail since the reports are the hypothetical top retrievals.

### Radiologist scoring criteria

In this work, we develop a scoring system for radiologists to evaluate the quality of candidate reports. The goals of our scoring system are to be objective, limit radiologist bias, and change linearly with report quality. To this end, scores are determined by counting the number of errors that candidate reports make where types of errors are broken down into six different categories. By explicitly defining each error category, we clarify what should be classified as an error. Following ACR’s RADPEER^37^ program for peer review, we differentiate between clinically significant and clinically insignificant errors. The detailed scoring criteria allows us to analyze report quality based on the accuracy of its findings and the clinical impact of its mistakes.

### Textual based and natural language generation performance metrics

In this study we make use of two natural language generation metrics: BLEU and BERTScore. The BLEU scores were computed as BLEU-2 bigrams with the fast_bleu library for parallel scoring. BERTScore uses the contextual embeddings from a BERT model to compute similarity of two text sequences. We used the bert_score library directly and used the baseline-scaled, “distilroberta-base” version of the model.

### Clinically aware performance metrics

In addition to traditional natural language generation metrics, we also investigated metrics that were designed to capture clinical information in radiology reports. Since radiology reports are a special form of structured text that communicates diagnostics information, their quality depends highly on the correctness of clinical objects and descriptions, which is not a focus of traditional natural language metrics. To address this gap, the CheXbert labeler (which is improved from the CheXpert labeler)^12,13^ and RadGraph^23^, were developed to parse radiology reports. We investigated whether they could be used as clinically aware metrics. We defined a metric as the similarity between CheXbert labeled vectors of the generated report and test report, which contain 14 labels corresponding to 13 common medical conditions and the no-finding observation. We used the implementation here: https://github.com/stanfordmlgroup/CheXbert. We proposed a novel metric as the overlap in parsed RadGraph graph structures: the RadGraph entity and relation F1 score. RadGraph is an approach for parsing radiology reports into knowledge graphs containing entities (nodes) and relations (edges), which can capture radiology concept dependencies and semantic meaning. We used the model checkpoint as provided here: https://physionet.org/content/radgraph/1.0.0/^26^ and inference code as provided here: https://github.com/dwadden/dygiepp^38^ to generate RadGraph entities and relations on generated and test reports.

### Retrieval-based metric-oracle models

To generate metric-oracle reports, the most immediate attempt is to adopt methods akin to those for multi-label classification tasks. Namely, we can curate a set of medical conditions and obtain radiologist annotations for each condition over a training set of reports. Then, we can train a classifier that outputs the likelihood of having each condition given an X-ray image, and proceed to select the corresponding report templates for conditions with high likelihood^14^. Some more nuanced approaches paraphrase the curated templates after selection^36^. The attempt at templating for report generation is well-grounded in abundant experience in multi-label image classification as well as its highly controlled output space. However, its flaw is also prominent, in that it is restricted to a manually curated predefined set of medical conditions and report templates. It does not generalize to unseen or complex conditions, express combinations of conditions, or capture uncertainty in diagnoses. The CheXbert labeler, for instance, can classify 13 conditions and the no-finding observation^13^. This set is representative of common medical observations but not comprehensive. Therefore, while we may define a larger set of conditions with the help of radiologists, manual curation and templating are nevertheless too inflexible for optimizing with respect to automated metrics. To generate reports of higher quality, we consider matching reports more closely onto test reports. We can do so by either generating new text from scratch or retrieving free text from an existing corpus of reports written by radiologists, given an X-ray image^33,35^. Out of the two approaches, retrieval-based methods have the advantage of a controlled output space that is the set of training report corpus, instead of an unpredictable output space produced by generation from scratch. Retrieval-based methods also improve upon templating, because the report corpus may capture the full set of real-world occurring conditions, combinations of conditions and uncertainty, if the training report corpus is representative of future reports to be written. Therefore, in this study, we use retrieval-based methods to generate metric-oracle reports.

### Statistical analysis

#### Metric-radiologist alignment

The alignment of metrics with radiologists’ scoring was determined using the Kendall tau-b correlation coefficient. We construct bootstrap confidence intervals by creating 1,000 resamples with replacement where each resample size is the number of studies (50). In this calculation, the number of errors is the mean number across all raters. Based on the presence/lack of overlap of the 95% bootstrap confidence intervals, we assert whether differences in Kendall tau values are statistically significant.

#### Metric failure modes

We conduct one-sided two-sample t tests on pairs of metrics’ error counts for total errors and clinically significant errors within each of the six error categories. We assume equal population variances for the t tests. We take the error count of one radiologist and one study as one data point. Because there are six radiologists and 50 studies, we have 300 data points per metric for either total errors or clinically significant errors and for one error category. With 4 metrics, there are 12 unique pairs of two different metrics for one-sided two-sample t tests with (300 + 300 - 2 = 598) degrees of freedom. We use the Benjamini-Hochberg Procedure with a False Discovery Rate (FDR) of 1% to account for multiple-hypothesis testing on 12 tests within an error type and an error category, and determine the significance of a metric having a more/less prominent failure mode compared with other metrics.

#### Prior models evaluation

To evaluate performance of metric-oracle models and prior state-of-the-art models, we construct bootstrap confidence intervals by taking 5,000 resamples with replacement of metric scores assigned to generated reports. Based on the presence/lack of overlap of the 95% bootstrap confidence intervals, we assert whether a model’s performance is statistically significantly better than another’s.

#### Composite metric RadCliQ

The linear regression model used to predict the total number of errors was evaluated using the Kendall-tau b statistical test. This test produces a tau-value correlation coefficient and a corresponding p-value which was used to determine the significance of the result (p-value < 0.01). The analyses were performed using statsmodels, scikit-learn and SciPy packages in Python.

### RadGraph metric-oracle model entities and relations match

The RadGraph F1 metric-oracle model retrieves reports with the highest F1 score match in terms of entities and relations. Specifically, we treat two entities as matched, if their tokens (words in the original report) and labels (entity type) match. We treat two relations as matched, if their start and end entities match and the relation type matches. These criteria are consistent with what the RadGraph authors have done. For combining entities and relations, we take the average of F1 score of entity match and relation match respectively. We generated RadGraph entities and relations for each report in the training and test corpora. We implemented the metric-oracle model by finding, for each report in the test set, which report in the training set is the best match based on the average of entity and relation F1 scores. For reports without nonzero F1 score matches, we used the most frequent report in the training set, “*No acute cardiopulmonary process*,” as the metric-oracle report in the radiologist experiment.

### Implementation of prior report generation methods

We used the following implementations of prior methods in radiology report generation: M^2^ Trans: https://github.com/ysmiura/ifcc^15,38^. R2Gen: https://github.com/cuhksz-nlp/R2Gen^16^. CXR-RePaiR: https://github.com/rajpurkarlab/CXR-RePaiR^17^. WCL: https://github.com/zzxslp/WCL^18^. CvT2DistilGPT2: https://github.com/aehrc/cvt2distilgpt2^19^. For each study ID, if the model generated multiple reports corresponding to different X-ray images for the same study, we used the generated report corresponding to the anterior-posterior (AP) or posterior-anterior (PA) view if any was present. If both were present, we randomly chose a report out of the two. If neither was present, we randomly chose a report out of the available reports corresponding to other views. Among variations of CXR-RePaiR, we chose CXR-RePaiR-2 to be consistent with their original study^17^.

## Data Availability

The data used in the study is available with credentialed access at: https://physionet.org/content/mimic-cxr-jpg/2.0.0/. Credentialed access can be obtained via an application to PhysioNet.

## Code Availability

The code for computing the composite metric RadCliQ and individual metrics is made publicly available at: https://drive.google.com/drive/folders/1Fe81n9IMZpc4y99K-7c5aGxPNdiij7NS?usp=sharing.

## Acknowledgements

We thank M.A. Endo MD for helpful review and feedback on the radiologist evaluation survey design and the manuscript. Support for this work was provided in part by the Medical Imaging Data Resource Center (MIDRC) under contracts 75N92020C00008 and 75N92020C00021 from the National Institute of Biomedical Imaging and Bioengineering (NIBIB) of the National Institutes of Health.

## Contributions

F.Y., M.E. and R.K. contributed equally to the design, implementation and analyses of all aspects of this study. I.P., A.T., E.P.R., E.K.U.N.F., H.M.H.L. and V.K.V. provided suggestions on the setup of the radiologist evaluation survey and provided annotations in the radiologist evaluation process. Z.S.H.A. contributed to the design of the illustrations and figures. A.Y.N., C.P.L., V.K.V. and P.R. oversaw and provided guidance on the study. All authors approved the final version.

## Corresponding author

Correspondence to Pranav Rajpurkar, PhD.

## Extended data

### Supplementary information

**Supplementary Fig. 1:**
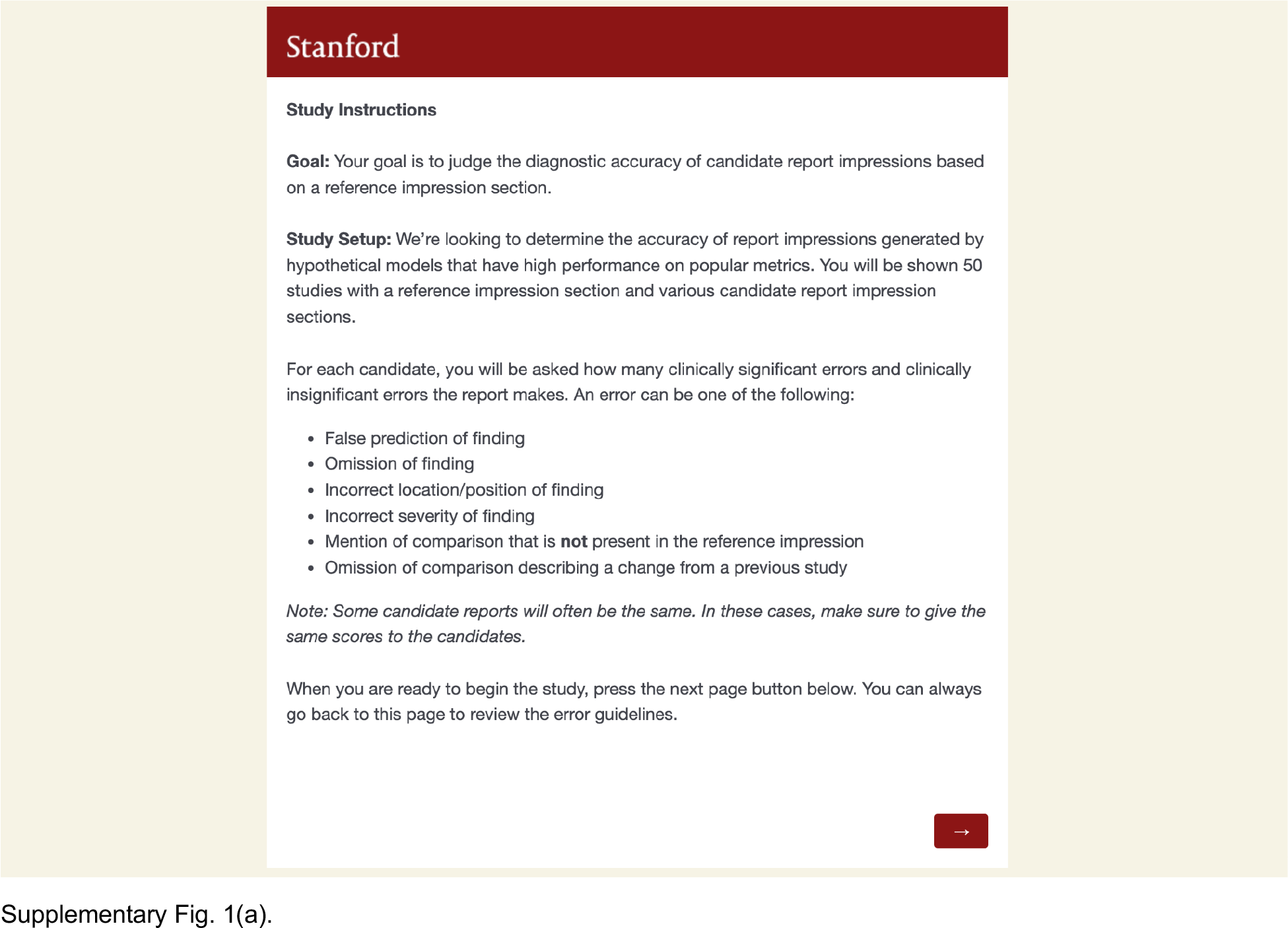
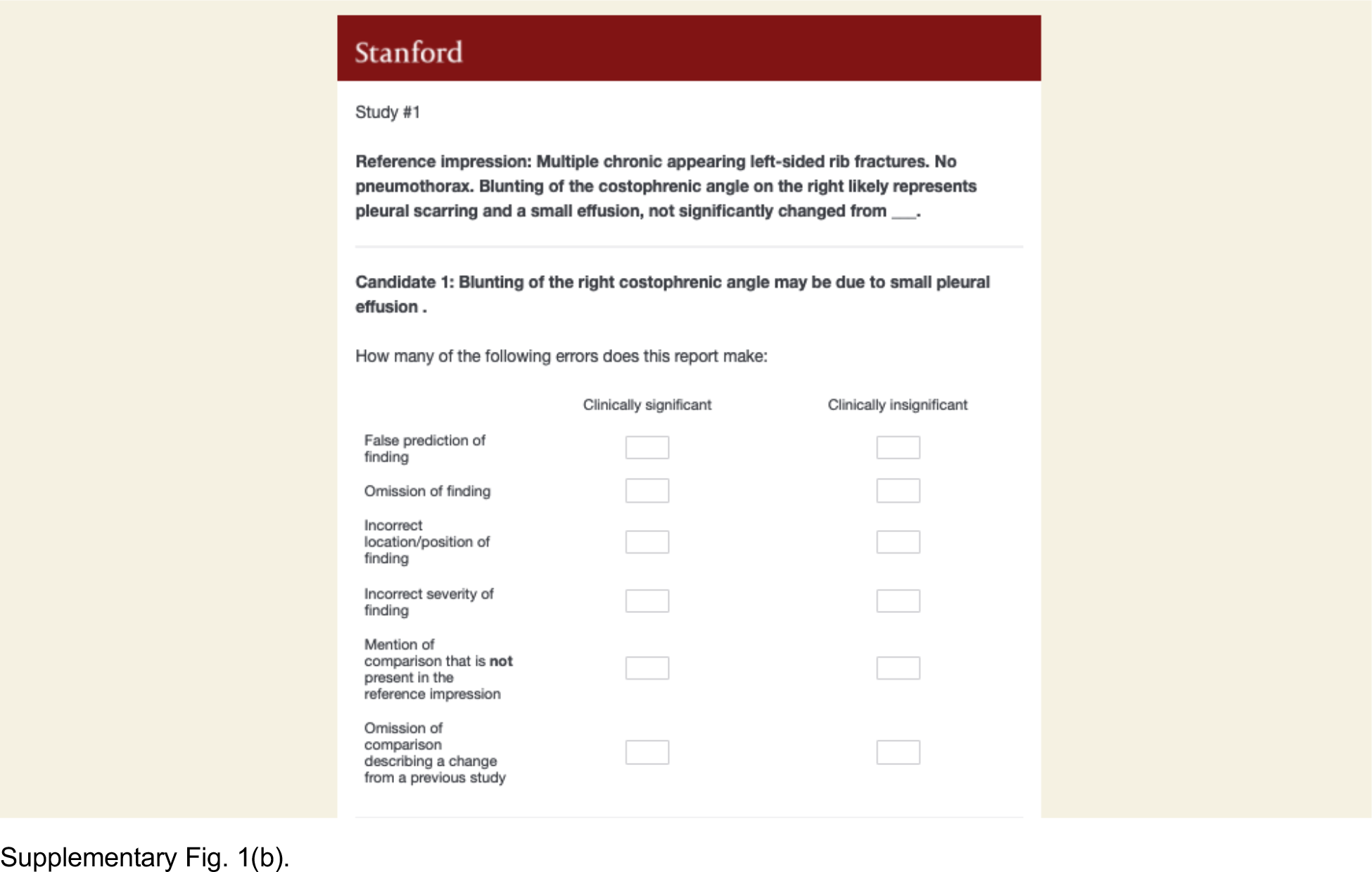
Radiologist survey interface and example question. **a**, Radiologist evaluation survey instructions and interface on Qualtrics. **b**, Interface for evaluating a pair of a test report (denoted as “Reference Impression”) and a metric-oracle report (denoted as “Candidate 1”). The survey asks radiologists to input the number of clinically significant and insignificant errors for six error categories.

**Supplementary Fig. 2:**
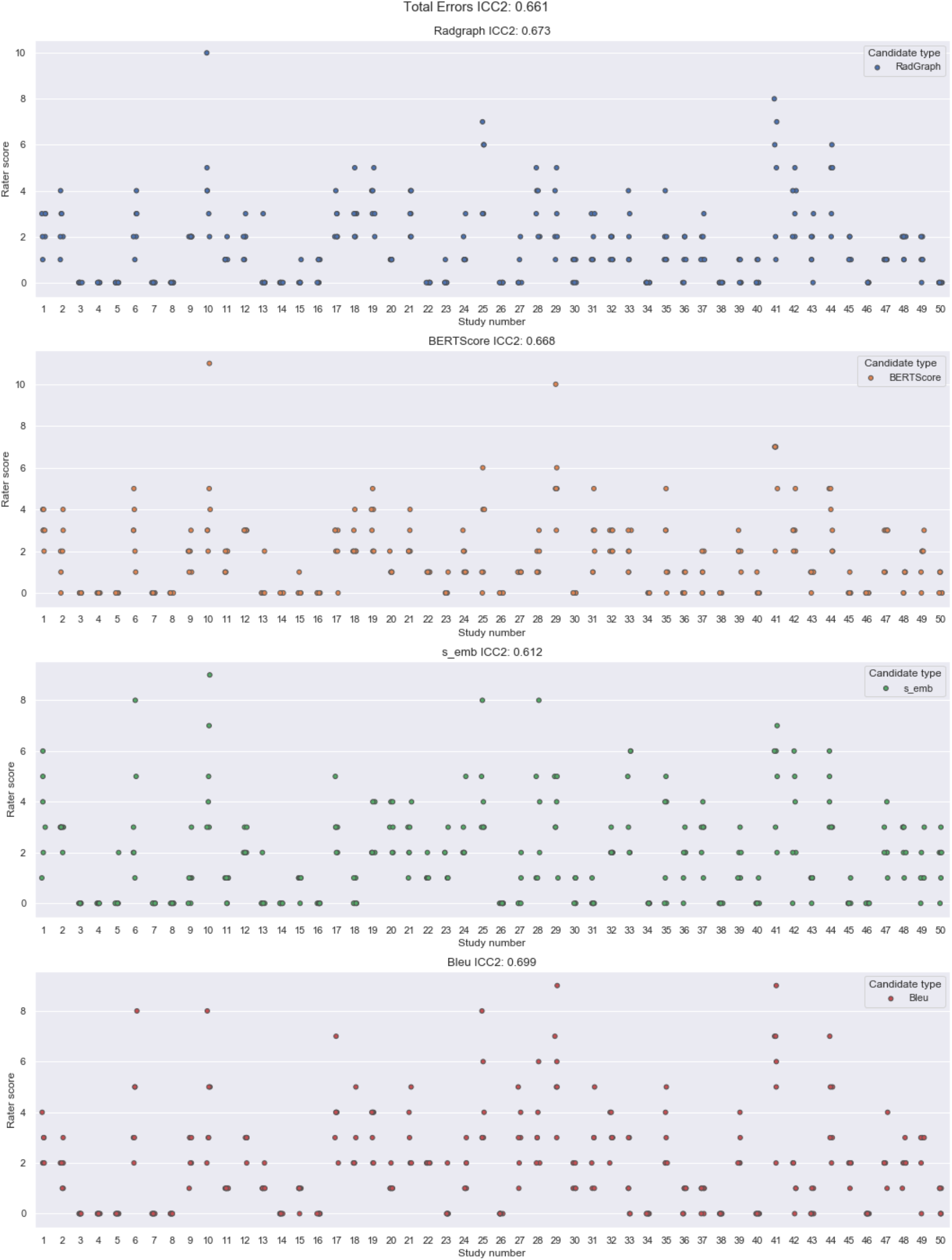
Dotplot of the radiologist total error scores on the 50 studies and corresponding intraclass correlation. Candidate scores are split up by metric-oracle method. Each dot represents a single radiologist’s score for a candidate report.

**Supplementary Table 1:**
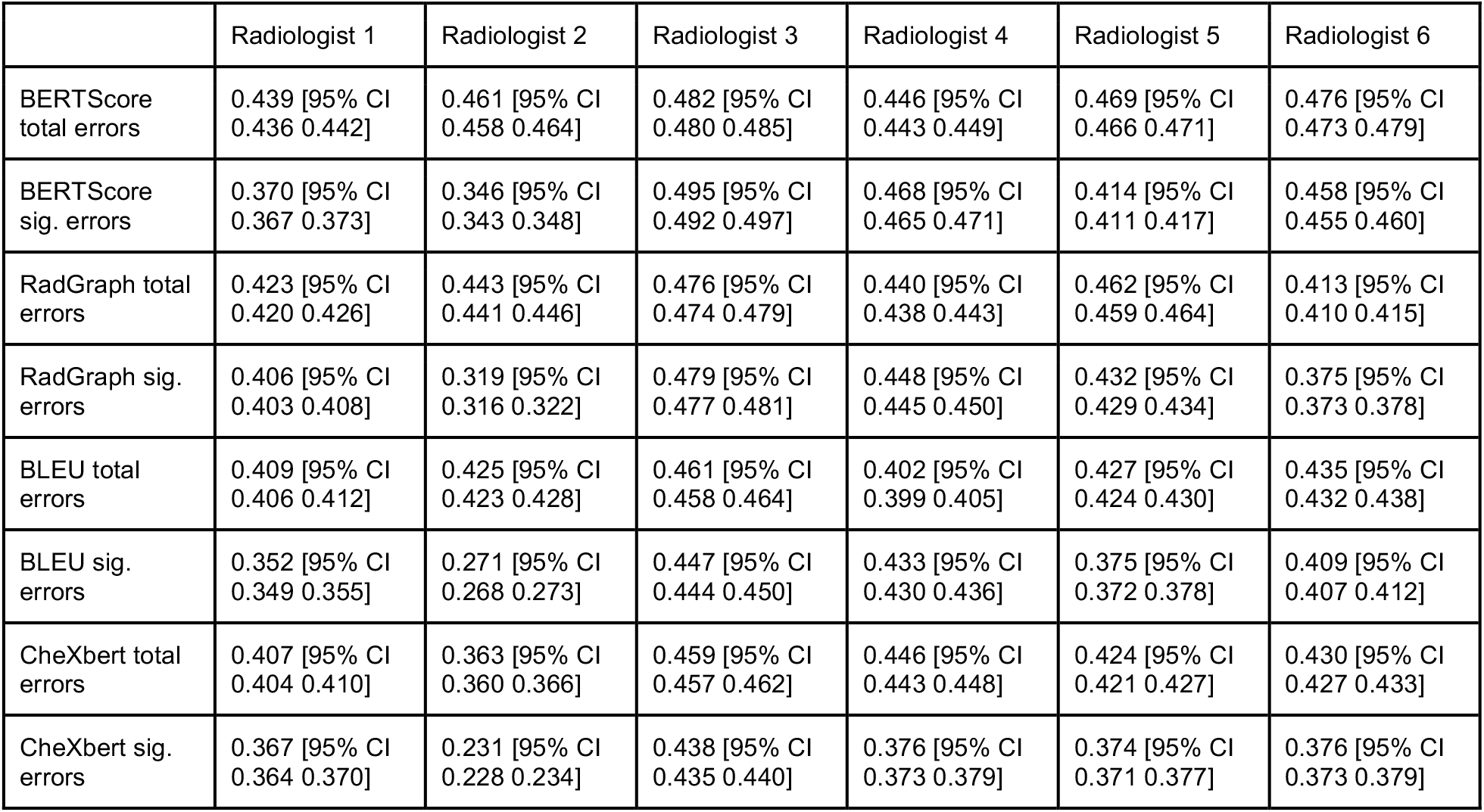
Per-radiologist Kendall rank correlation coefficient (tau-b) values quantifying metric-radiologist alignment.

|  | Radiologist 1 | Radiologist 2 | Radiologist 3 | Radiologist 4 | Radiologist 5 | Radiologist 6 |
| --- | --- | --- | --- | --- | --- | --- |
| BERTScore total errors | 0.439 [95% CI 0.436 0.442] | 0.461 [95% CI 0.458 0.464] | 0.482 [95% CI 0.480 0.485] | 0.446 [95% CI 0.443 0.449] | 0.469 [95% CI 0.466 0.471] | 0.476 [95% CI 0.473 0.479] |
| BERTScore sig. errors | 0.370 [95% CI 0.367 0.373] | 0.346 [95% CI 0.343 0.348] | 0.495 [95% CI 0.492 0.497] | 0.468 [95% CI 0.465 0.471] | 0.414 [95% CI 0.411 0.417] | 0.458 [95% CI 0.455 0.460] |
| RadGraph total errors | 0.423 [95% CI 0.420 0.426] | 0.443 [95% CI 0.441 0.446] | 0.476 [95% CI 0.474 0.479] | 0.440 [95% CI 0.438 0.443] | 0.462 [95% CI 0.459 0.464] | 0.413 [95% CI 0.410 0.415] |
| RadGraph sig. errors | 0.406 [95% CI 0.403 0.408] | 0.319 [95% CI 0.316 0.322] | 0.479 [95% CI 0.477 0.481] | 0.448 [95% CI 0.445 0.450] | 0.432 [95% CI 0.429 0.434] | 0.375 [95% CI 0.373 0.378] |
| BLEU total errors | 0.409 [95% CI 0.406 0.412] | 0.425 [95% CI 0.423 0.428] | 0.461 [95% CI 0.458 0.464] | 0.402 [95% CI 0.399 0.405] | 0.427 [95% CI 0.424 0.430] | 0.435 [95% CI 0.432 0.438] |
| BLEU sig. errors | 0.352 [95% CI 0.349 0.355] | 0.271 [95% CI 0.268 0.273] | 0.447 [95% CI 0.444 0.450] | 0.433 [95% CI 0.430 0.436] | 0.375 [95% CI 0.372 0.378] | 0.409 [95% CI 0.407 0.412] |
| CheXbert total errors | 0.407 [95% CI 0.404 0.410] | 0.363 [95% CI 0.360 0.366] | 0.459 [95% CI 0.457 0.462] | 0.446 [95% CI 0.443 0.448] | 0.424 [95% CI 0.421 0.427] | 0.430 [95% CI 0.427 0.433] |
| CheXbert sig. errors | 0.367 [95% CI 0.364 0.370] | 0.231 [95% CI 0.228 0.234] | 0.438 [95% CI 0.435 0.440] | 0.376 [95% CI 0.373 0.379] | 0.374 [95% CI 0.371 0.377] | 0.376 [95% CI 0.373 0.379] |

**Supplementary Table 2(a):** Radiologist evaluation of metric-oracles in terms of total number of errors in six error categories, averaged over 6 radiologists and 50 studies.

|  | Error 1 | Error 2 | Error 3 | Error 4 | Error 5 | Error 6 | Total |
| --- | --- | --- | --- | --- | --- | --- | --- |
| BLEU | 0.807 | 0.550 | 0.113 | 0.133 | 0.140 | 0.097 | 1.840 |
| CheXbert | 0.597 | 0.443 | 0.227 | 0.197 | 0.150 | 0.093 | 1.707 |
| BERTScore | 0.477 | 0.523 | 0.183 | 0.153 | 0.077 | 0.113 | 1.527 |
| RadGraph | 0.427 | 0.573 | 0.147 | 0.160 | 0.077 | 0.110 | 1.493 |

**Supplementary Table 2(b):** Radiologist evaluation of metric-oracles in terms of number of clinically significant errors in six error categories, averaged over 6 radiologists and 50 studies.

|  | Error 1 | Error 2 | Error 3 | Error 4 | Error 5 | Error 6 | Total |
| --- | --- | --- | --- | --- | --- | --- | --- |
| BLEU | 0.607 | 0.353 | 0.087 | 0.093 | 0.107 | 0.077 | 1.323 |
| CheXbert | 0.430 | 0.263 | 0.193 | 0.176 | 0.107 | 0.077 | 1.247 |
| BERTScore | 0.363 | 0.310 | 0.147 | 0.117 | 0.053 | 0.083 | 1.073 |
| RadGraph | 0.300 | 0.343 | 0.133 | 0.143 | 0.053 | 0.070 | 1.043 |

**Supplementary Table 3(a):** Significance of BLEU having a *more prominent failure mode* than BERTScore and RadGraph F1 in terms of *total errors* in *false prediction of finding*, as determined by the Benjamini-Hochberg Procedure with False Discovery Rate (FDR) of 1%.

|  | One-sided two-sample t test<br>p-value | Benjamini-Hochberg<br>Procedure critical value with<br>False Discovery Rate (FDR)<br>of 1% | Whether result is significant |
| --- | --- | --- | --- |
| BLEU > CheXbert | 3.79e-3 | 2.50e-3 | N |
| BLEU > BERTScore | 9.50e-6 | 1.67e-3 | Y |
| BLEU > RadGraph | 1.07e-7 | 8.33e-4 | Y |
| CheXbert > BLEU | 9.96e-1 | 8.33e-3 | N |
| CheXbert > BERTScore | 7.65e-2 | 4.17e-3 | N |
| CheXbert > RadGraph | 6.39e-3 | 3.33e-3 | N |
| BERTScore > BLEU | 1.00e0 | 9.17e-3 | N |
| BERTScore > CheXbert | 9.51e-1 | 6.67e-3 | N |
| BERTScore > RadGraph | 2.24e-1 | 5.00e-3 | N |
| RadGraph > BLEU | 1.00e0 | 1.00e-2 | N |
| RadGraph > CheXbert | 9.94e-1 | 7.50e-3 | N |
| RadGraph > BERTScore | 7.76e-1 | 5.83e-3 | N |

**Supplementary Table 3(b):** Significance of BLEU having a *more prominent failure mode* than BERTScore and RadGraph F1 in terms of *clinically significant errors* in *false prediction of finding*, as determined by the Benjamini-Hochberg Procedure with False Discovery Rate (FDR) of 1%.

|  | One-sided two-sample t test<br>p-value | Benjamini-Hochberg<br>Procedure critical value with<br>False Discovery Rate (FDR)<br>of 1% | Whether result is significant |
| --- | --- | --- | --- |
| BLEU > CheXbert | 3.68e-3 | 2.50e-3 | N |
| BLEU > BERTScore | 1.48e-4 | 1.67e-3 | Y |
| BLEU > RadGraph | 6.44e-7 | 8.33e-4 | Y |
| CheXbert > BLEU | 9.96e-1 | 8.33e-3 | N |
| CheXbert > BERTScore | 1.34e-1 | 5.00e-3 | N |
| CheXbert > RadGraph | 9.77e-3 | 3.33e-3 | N |
| BERTScore > BLEU | 1.00e0 | 9.17e-3 | N |
| BERTScore > CheXbert | 8.66e-1 | 5.83e-3 | N |
| BERTScore > RadGraph | 1.33e-1 | 4.17e-3 | N |
| RadGraph > BLEU | 1.00e0 | 1.00e-2 | N |
| RadGraph > CheXbert | 9.90e-1 | 7.50e-3 | N |
| RadGraph > BERTScore | 8.67e-1 | 6.67e-3 | N |

**Supplementary Table 3(c):** Significance of BLEU having a *less prominent failure mode* than CheXbert vector similarity in terms of *total errors* in *incorrect location/position of finding*, as determined by the Benjamini-Hochberg Procedure with False Discovery Rate (FDR) of 1%.

|  | One-sided two-sample t test<br>p-value | Benjamini-Hochberg<br>Procedure critical value with<br>False Discovery Rate (FDR)<br>of 1% | Whether result is significant |
| --- | --- | --- | --- |
| BLEU < CheXbert | 4.83e-4 | 8.33e-4 | Y |
| BLEU < BERTScore | 1.60e-2 | 2.50e-3 | N |
| BLEU < RadGraph | 1.51e-1 | 5.00e-3 | N |
| CheXbert < BLEU | 1.00e0 | 1.00e-2 | N |
| CheXbert < BERTScore | 8.90e-1 | 7.50e-3 | N |
| CheXbert < RadGraph | 9.89e-1 | 9.17e-3 | N |
| BERTScore < BLEU | 9.84e-1 | 8.33e-3 | N |
| BERTScore < CheXbert | 1.10e-1 | 3.33e-3 | N |
| BERTScore < RadGraph | 8.63e-1 | 6.67e-3 | N |
| RadGraph < BLEU | 8.49e-1 | 5.83e-3 | N |
| RadGraph < CheXbert | 1.14e-2 | 1.67e-3 | N |
| RadGraph < BERTScore | 1.37e-1 | 4.17e-3 | N |

**Supplementary Table 3(d):**
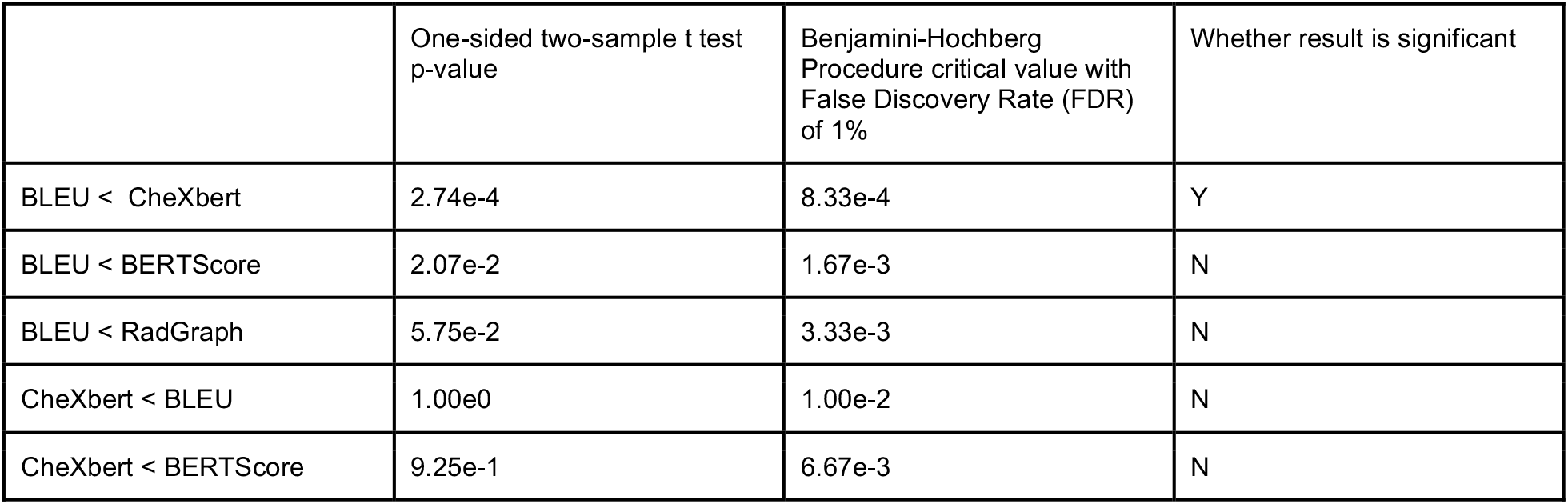
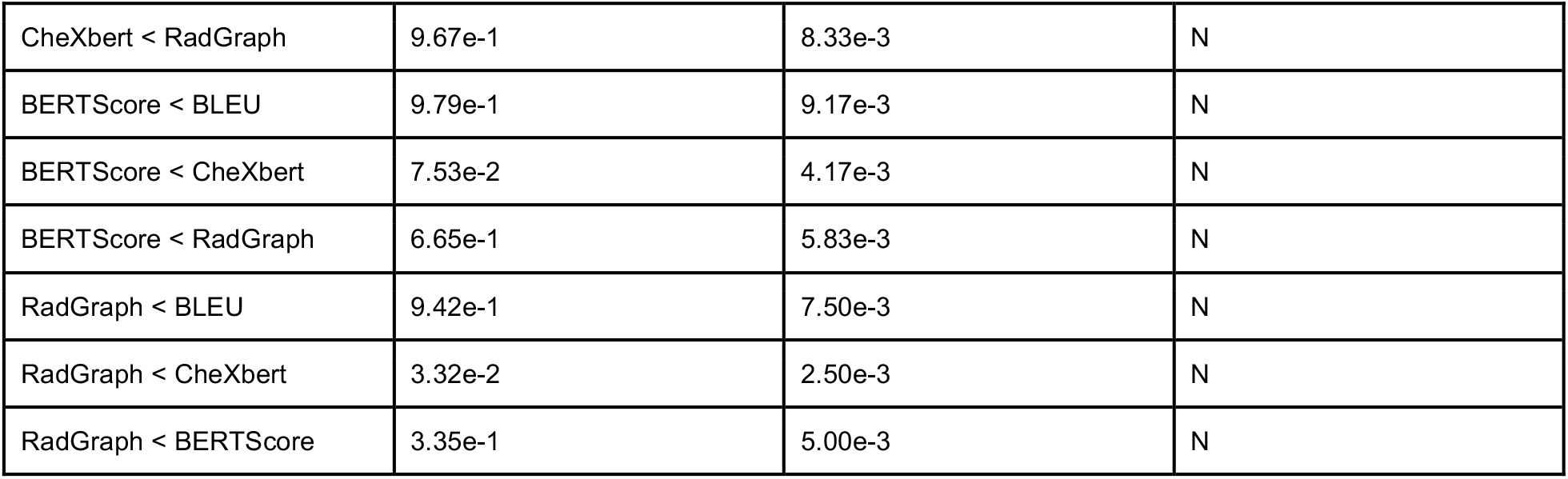
Significance of BLEU having a *less prominent failure mode* than CheXbert vector similarity in terms of *clinically significant errors* in *incorrect location/position of finding*, as determined by the Benjamini-Hochberg Procedure with False Discovery Rate (FDR) of 1%.

**Supplementary Table 4:** Average metric scores of metric-oracle models and prior models, as the average, lower-bound, and upper-bound of the 95% confidence interval.

|  | Avg BLEU | Avg BERTScore | Avg CheXbert | Avg RadGraph |
| --- | --- | --- | --- | --- |
| Metric-oracle model | 0.566 [95% CI 0.566 0.566] | 0.729 [95% CI 0.729 0.729] | 0.956 [95% CI 0.956 0.956] | 0.677 [95% CI 0.677 0.678] |
| Random | 0.048 [95% CI 0.048 0.049] | 0.214 [95% CI 0.214 0.214] | 0.280 [95% CI 0.280 0.280] | 0.049 [95% CI 0.049 0.049] |
| M<sup>2</sup> Trans | 0.087 [95% CI 0.087 0.087] | 0.227 [95% CI 0.227 0.227] | 0.268 [95% CI 0.268 0.268] | 0.110 [95% CI 0.110 0.111] |
| R2Gen | 0.059 [95% CI 0.059 0.059] | 0.186 [95% CI 0.186 0.186] | 0.204 [95% CI 0.203 0.204] | 0.057 [95% CI 0.057 0.057] |
| CXR-RePaiR | 0.055 [95% CI 0.055 0.055] | 0.191 [95% CI 0.191 0.191] | 0.379 [95% CI 0.379 0.379] | 0.091 [95% CI 0.090 0.091] |
| WCL | 0.064 [95% CI 0.064 0.064] | 0.188 [95% CI 0.188 0.189] | 0.218 [95% CI 0.218 0.218] | 0.068 [95% CI 0.068 0.068] |
| CvT2DistilGPT2 | 0.066 [95% CI 0.066 0.066] | 0.192 [95% CI 0.192 0.193] | 0.264 [95% CI 0.264 0.264] | 0.073 [95% CI 0.073 0.073] |

**Supplementary Table 5:**
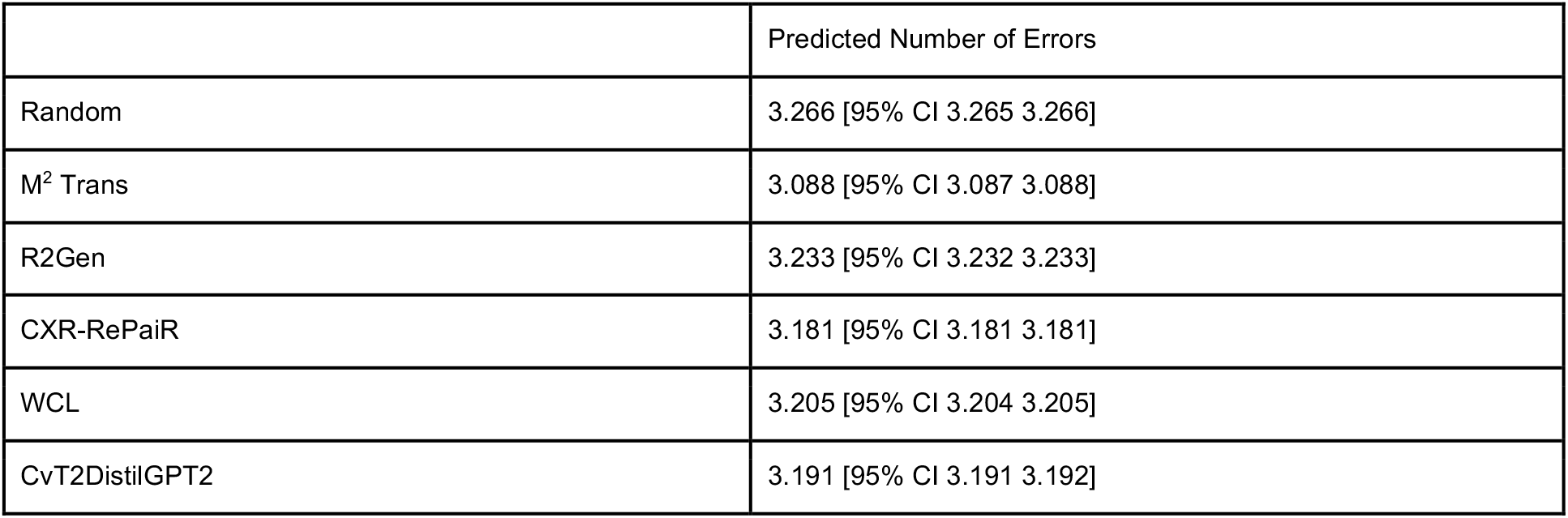
Predicted total number of errors of prior models, as the average, lower-bound, and upper-bound of the 95% confidence interval.

|  | Predicted Number of Errors |
| --- | --- |
| Random | 3.266 [95% CI 3.265 3.266] |
| M<sup>2</sup> Trans | 3.088 [95% CI 3.087 3.088] |
| R2Gen | 3.233 [95% CI 3.232 3.233] |
| CXR-RePaiR | 3.181 [95% CI 3.181 3.181] |
| WCL | 3.205 [95% CI 3.204 3.205] |
| CvT2DistilGPT2 | 3.191 [95% CI 3.191 3.192] |

## References

1. Silver, D. et al. Mastering the game of Go with deep neural networks and tree search. Nature 529, 484–489 (2016).

2. Silver, D. et al. Mastering the game of Go without human knowledge. Nature 550, 354–359 (2017).

3. Silver, D. et al. A general reinforcement learning algorithm that masters chess, shogi, and Go through self-play. Science 362, 1140–1144 (2018).

4. Schrittwieser, J. et al. Mastering Atari, Go, chess and shogi by planning with a learned model. Nature 588, 604–609 (2020).

5. Li, Y. et al. Competition-Level Code Generation with AlphaCode. (2022) doi:10.48550/arXiv.2203.07814.

6. Chen, M. et al. Evaluating Large Language Models Trained on Code. (2021) doi:10.48550/arXiv.2107.03374.

7. Bojarski, M. et al. End to End Learning for Self-Driving Cars. (2016) doi:10.48550/arXiv.1604.07316.

8. Fridman, L. et al. MIT Advanced Vehicle Technology Study: Large-Scale Naturalistic Driving Study of Driver Behavior and Interaction with Automation. (2017) doi:10.1109/ACCESS.2019.2926040.

9. Rajpurkar, P., Chen, E., Banerjee, O. & Topol, E. J. AI in health and medicine. Nat. Med. 28, 31–38 (2022).

10. Senior, A. W. et al. Improved protein structure prediction using potentials from deep learning. Nature 577, 706–710 (2020).

11. Jumper, J. et al. Highly accurate protein structure prediction with AlphaFold. Nature 596, 583–589 (2021).

12. Irvin, J. et al. CheXpert: A Large Chest Radiograph Dataset with Uncertainty Labels and Expert Comparison. (2019) doi:10.48550/arXiv.1901.07031.

13. Smit, A. et al. CheXbert: Combining Automatic Labelers and Expert Annotations for Accurate Radiology Report Labeling Using BERT. (2020) doi:10.48550/arXiv.2004.09167.

14. Pino, P., Parra, D., Besa, C. & Lagos, C. Clinically Correct Report Generation from Chest X-Rays Using Templates. in Machine Learning in Medical Imaging 654–663 (Springer, Cham, 2021).

15. Miura, Y., Zhang, Y., Tsai, E. B., Langlotz, C. P. & Jurafsky, D. Improving Factual Completeness and Consistency of Image-to-Text Radiology Report Generation. (2020) doi:10.48550/arXiv.2010.10042.

16. Chen, Z., Song, Y., Chang, T.-H. & Wan, X. Generating Radiology Reports via Memory-driven Transformer. (2020) doi:10.48550/arXiv.2010.16056.

17. Endo, M., Krishnan, R., Krishna, V., Ng, A. Y. & Rajpurkar, P. Retrieval-Based Chest X-Ray Report Generation Using a Pre-trained Contrastive Language-Image Model. in Machine Learning for Health 209–219 (PMLR, 2021).

18. Yan, A. et al. Weakly Supervised Contrastive Learning for Chest X-Ray Report Generation. (2021) doi:10.48550/arXiv.2109.12242.

19. Nicolson, A., Dowling, J. & Koopman, B. Improving Chest X-Ray Report Generation by Leveraging Warm-Starting. (2022) doi:10.48550/arXiv.2201.09405.

20. Zhou, H.-Y. et al. Generalized radiograph representation learning via cross-supervision between images and free-text radiology reports. Nature Machine Intelligence 4, 32–40 (2022).

21. Hossain, M. Z., Sohel, F., Shiratuddin, M. F. & Laga, H. A Comprehensive Survey of Deep Learning for Image Captioning. (2018) doi:10.48550/arXiv.1810.04020.

22. William Boag MIT, U. S. A., Hassan Kané WL Research, USA, Saumya Rawat MIT, U. S. A., Jesse Wei Beth Israel Deaconess Medical Center, Department of Radiology, USA & Alexander Goehler Beth Israel Deaconess Medical Center, Department of Radiology, USA. A Pilot Study in Surveying Clinical Judgments to Evaluate Radiology Report Generation. ACM Conferences https://dl.acm.org/doi/abs/10.1145/3442188.3445909.

23. Jain, S. et al. RadGraph: Extracting Clinical Entities and Relations from Radiology Reports. (2021) doi:10.48550/arXiv.2106.14463.

24. Johnson, A. E. W. et al. MIMIC-CXR, a de-identified publicly available database of chest radiographs with free-text reports. Scientific Data 6, 1–8 (2019).

25. Johnson, A. E. W. et al. MIMIC-CXR-JPG, a large publicly available database of labeled chest radiographs. (2019) doi:10.48550/arXiv.1901.07042.

26. Goldberger, A. L. et al. PhysioBank, PhysioToolkit, and PhysioNet. Circulation vol. 101 (2000).

27. Kishore Papineni IBM T. J. Watson Research Center, Yorktown Heights, N., Salim Roukos IBM T. J. Watson Research Center, Yorktown Heights, N., Todd Ward IBM T. J. Watson Research Center, Yorktown Heights, N. & Wei-Jing Zhu IBM T. J. Watson Research Center, Yorktown Heights, NY. Bleu. DL Hosted proceedings https://dl.acm.org/doi/abs/10.3115/1073083.1073135.

28. Zhang, T., Kishore, V., Wu, F., Weinberger, K. Q. & Artzi, Y. BERTScore: Evaluating Text Generation with BERT. (2019) doi:10.48550/arXiv.1904.09675.

29. Vedantam, R., Zitnick, C. L. & Parikh, D. CIDEr: Consensus-based Image Description Evaluation. (2014) doi:10.48550/arXiv.1411.5726.

30. Alon Lavie Carnegie Mellon University, Pittsburgh, P. & Abhaya Agarwal Carnegie Mellon University, Pittsburgh, PA. Meteor. DL Hosted proceedings https://dl.acm.org/doi/abs/10.5555/1626355.1626389.

31. Lin, C.-Y. ROUGE: A Package for Automatic Evaluation of Summaries. in Text Summarization Branches Out 74–81 (2004).

32. Anderson, P., Fernando, B., Johnson, M. & Gould, S. SPICE: Semantic Propositional Image Caption Evaluation. (2016) doi:10.48550/arXiv.1607.08822.

33. Deep learning in generating radiology reports: A survey. Artif. Intell. Med. 106, 101878 (2020).

34. Zhou, Y., Huang, L., Zhou, T., Fu, H. & Shao, L. Visual-Textual Attentive Semantic Consistency for Medical Report Generation. in Proceedings of the IEEE/CVF International Conference on Computer Vision 3985–3994 (2021).

35. Wang, X., Zhang, Y., Guo, Z. & Li, J. ImageSem at ImageCLEF 2018 Caption Task: Image Retrieval and Transfer Learning. (2018).

36. Li, C. Y., Liang, X., Hu, Z. & Xing, E. P. Knowledge-Driven Encode, Retrieve, Paraphrase for Medical Image Report Generation. AAAI 33, 6666–6673 (2019).

37. Goldberg-Stein, S. et al. ACR RADPEER Committee White Paper with 2016 Updates: Revised Scoring System, New Classifications, Self-Review, and Subspecialized Reports. J. Am. Coll. Radiol. 14, 1080–1086 (2017).

38. Wadden, D., Wennberg, U., Luan, Y. & Hajishirzi, H. Entity, Relation, and Event Extraction with Contextualized Span Representations. (2019).

